# Longitudinal Characterization of Nociplasticity in Systemic Lupus Erythematosus: A Nationwide Registry Study

**DOI:** 10.64898/2026.08.20.26360943

**Authors:** Cho-Yi Huang, Christophe Tanguay-Sabourin, Yuhan Liu, Sofia Pedro, Troy C. Dildine, Selen Bozkurt, Patricia Katz, Kaleb Michaud, Titilola Falasinnu

## Abstract

**Background:** Nociplasticity is common in systemic lupus erythematosus (SLE), yet its longitudinal trajectory remain poorly characterized.

**Methods:** Patients with SLE in the FORWARD Databank were classified as Minimal, Type 1, Type 2, or Mixed using the Polysymptomatic Distress Scale (PSD≥8) and the Systemic Lupus Activity Questionnaire (SLAQ) inflammatory domain score (≥2). Cross-sectional analyses (N=372) compared pain, function, disability, depression, organ damage, and medication use. Longitudinal analyses (n=301; median 3.7 years) characterized phenotype transitions using continuous-time Markov models and identified latent trajectory subgroups using joint group-based trajectory modeling (GBTM).

**Findings:** At baseline, 29% were Minimal, 12% Type 1, 13% Type 2, and 47% Mixed. Functional impairment increased stepwise: SF-36 PCS worsened from 49.7 in Minimal to 30.0 in Mixed; PROMIS Pain Interference from 46.2 to 63.6 (both p<0.001). BILD organ damage was highest in Mixed (3.9 vs 2.6–2.9; p<0.001). Depression rose from 3% to 43% and opioid use reached 44% in Mixed. Longitudinally, Minimal and Mixed were persistent (mean duration 2.0 and 1.8 years; one-year retention 70%), while Type 1 and Type 2 were transient (∼0.5 years; retention 18% and 28%). Exit trajectories were asymmetric: Type 1 moved preferentially to Minimal (49% of exits), whereas Type 2 moved to Mixed (65%; p<0.001). Population-average PSD was nearly flat (+0.014 SD/year, p=0.07); opioid use declined to near zero in Minimal and Type 1 but remained high in Type 2 and Mixed. Joint GBTM identified four severity classes along a Minimal-to-Mixed diagonal, indicating that the two axes scale together longitudinally and that pure Type 1 and Type 2 are transient.

**Interpretation:** Nociplastic phenotypes in SLE are persistent, severity-stratified, and associated with functional impairment, depression, cumulative damage, and sustained opioid burden. Type 1 and Type 2 are transient states with divergent exits: the former resolving, the latter progressing.

**Funding:** Lupus Research Alliance

**Research in context:** *Evidence before this study:* The Type 1 and Type 2 systemic lupus erythematosus (SLE) framework distinguishes inflammatory manifestations from symptoms associated with nociplastic pain and has provided a clinically meaningful approach to understanding the heterogeneity of SLE. Patient-reported instruments, including the polysymptomatic distress (PSD) scale combined with inflammatory symptom assessment, have enabled classification of these phenotypes in both research and clinical settings. A search of PubMed, Embase, and Web of Science (from database inception to June 01, 2026) using the terms “systemic lupus erythematosus”, “Type 1 SLE”, “Type 2 SLE”, “nociplastic pain”, “nociplasticity”, “polysymptomatic distress”, “trajectory”, and “transition”, together with screening of reference lists of relevant articles, identified studies describing cross-sectional phenotype distributions, intermittent and persistent Type 2 symptoms, and molecular differences between phenotypes. However, whether Type 1 and Type 2 phenotypes represent stable disease states or transient manifestations of a patient’s underlying longitudinal symptom trajectory is unknown.

*Added value of this study:* This study analyzed prospectively collected patient-reported data from 301 SLE patients enrolled in the nationwide FORWARD registry over a median follow-up of 3.7 years to characterize the longitudinal behavior of Type 1 and Type 2 phenotypes. By combining continuous-time Markov modelling with group-based trajectory modelling, we distinguished threshold-defined phenotype transitions from underlying symptom trajectories. We found that Minimal and Mixed phenotypes were highly stable over time, whereas Type 1 and Type 2 phenotypes represented transient states with distinct transition patterns. Data-driven trajectories formed a stable severity continuum from Minimal to Mixed rather than persistent Type 1 and Type 2 classes. Higher nociplastic burden was also associated with sustained impairment in physical function, depression, medication burden, and opioid use.

*Implication of all the available evidence:* These findings suggest that nociplastic symptom burden represents a persistent and clinically important component of SLE that is not fully captured by inflammatory disease activity alone. Routine assessment of nociplastic symptoms alongside inflammatory activity could improve longitudinal patient stratification and identify individuals who may benefit from combined immunomodulatory and symptom-directed management. Future studies should determine whether phenotype-informed treatment strategies improve long-term patient outcomes.

## INTRODUCTION

Systemic lupus erythematosus (SLE) management has focused predominantly on controlling inflammatory disease activity, yet pain, fatigue, and other debilitating symptoms may remain substantial even when objective disease activity is controlled.^1^ These symptoms substantially impair quality of life, daily function, and workforce participation.^2,3^ Disease activity and organ damage explain only one-third of pain variance in SLE, implicating non-inflammatory mechanisms.^4^ The concept of nociplastic pain, arising from altered central nervous system pain modulation without clear peripheral tissue damage,^5^ offers a mechanistic framework for this disconnect.

Pisetsky et al.^6,7^ proposed a Type 1 and Type 2 SLE framework, wherein Type 1 symptoms reflect inflammatory disease activity (e.g., joint swelling, rash, serositis) and Type 2 symptoms encompass nociplastic manifestations including diffuse pain, fatigue, sleep disturbance, mood disruption, and cognitive dysfunction.^6^ Building on this framework, a clinician-assessed cohort of 212 patients classified 49% as minimal activity, 30% as Type 1, 8% as Type 2, and 13% as mixed.^8^ A subsequent patient-reported outcome (PRO)-based algorithm using the Systemic Lupus Activity Questionnaire (SLAQ), Polysymptomatic Distress Scale (PSD), and Patient Health Questionnaire-2 (PHQ-2) achieved 83% classification accuracy against clinician evaluation.^9^ In parallel, qualitative interviews distinguished intermittent Type 2 patients (generally well when inflammatory disease was inactive) from persistent Type 2 patients with chronic nociplastic symptoms.^10,11^ Most recently, RNA sequencing of pure phenotypes revealed distinct transcriptomic signatures, with interferon and monocyte pathways enriched in Type 1 versus B cell and neuromuscular pathways in Type 2.^12^

Converging evidence further supports these mechanisms: fibromyalgia prevalence in SLE is 15.8% (3.7 times that of the general population);^13^ functional MRI demonstrates altered central pain processing;^14^ and passive transfer of IgG from fibromyalgia patients induces pain behavior in mice.^15^ Integrating the evidence, the same group proposed the term Lupus-Associated Nociplasticity (LAN)^16^ and identified several critical gaps:^7^ prior studies have been cross-sectional, single-center, and have not evaluated longitudinal phenotype stability, functional burden, or treatment patterns.

We address these gaps using FORWARD Databank,^17^ a nationwide, community-based US registry for rheumatic diseases. Our objectives were to: (1) operationalize the Type 1/Type 2 framework using PROs alone and evaluate concordance with fibromyalgia criteria; (2) quantify the functional impact and treatment associations of nociplastic phenotypes; (3) assess longitudinal phenotype stability up to 7 years; and (4) characterize within-patient trajectories. These findings would establish the clinical relevance of nociplastic phenotyping for SLE management and provide the longitudinal evidence to guide phenotype-targeted treatment strategies.

## METHODS

### Study Population and Data Source

The FORWARD Databank (previously known as The National Databank for Rheumatic Diseases), is a longitudinal US registry in which patient with rheumatic diseases complete semiannual questionnaires by mail or online.^17^ Patients are enrolled by their rheumatologists nationwide, capturing community-based care outside of academic centers. We included patients with physician-confirmed or self-reported SLE diagnosis. Because the Polysymptomatic Distress Scale (PSD) and Systemic Lupus Activity Questionnaire (SLAQ) were introduced into the FORWARD questionnaire in 2016, analyses were restricted to visits on or after January 1, 2016 with complete concurrent SLAQ and PSD. Of 3,088 SLE patients, 1,004 had at least one visit after 2016; of these, 372 (37%) had concurrent SLAQ and PSD data. Longitudinal analyses further required ≥2 qualifying visits (n=301, median follow-up 3.7 years). Patient flow, survey availability by calendar year, and follow-up duration are shown in Supplementary Figure 1.

### Operationalizing Type 1 and Type 2 SLE Activity

We operationalized the Type 1/Type 2 framework as a fully patient-reported outcome (PRO)-based two-axis classification to permit phenotype assignment at each survey visit. The Type 1 axis used nine SLAQ inflammatory symptom items grouped into arthritis, mucocutaneous, serositis, and fever domains, each scored 0-3. The inflammatory score was defined as the maximum score across the four inflammatory domains. High Type 1 activity was defined as an inflammatory score ≥ 2.

The Type 2 nociplastic axis used the PSD (range 0-31), which combines the Widespread Pain Index (WPI) and Symptom Severity Score (SSS). High Type 2 activity was defined as PSD ≥ 8, a threshold proposed to identify clinically meaningful nociplastic.^7^

Fatigue, cognitive difficulty, and abdominal pain appear in the broader SLAQ item pool but were excluded from the Type 1 inflammatory domains, minimizing construct overlap between axes. Patients were classified as Minimal, Type 1, Type 2, or Mixed (Figure 1A).

**Figure 1.**
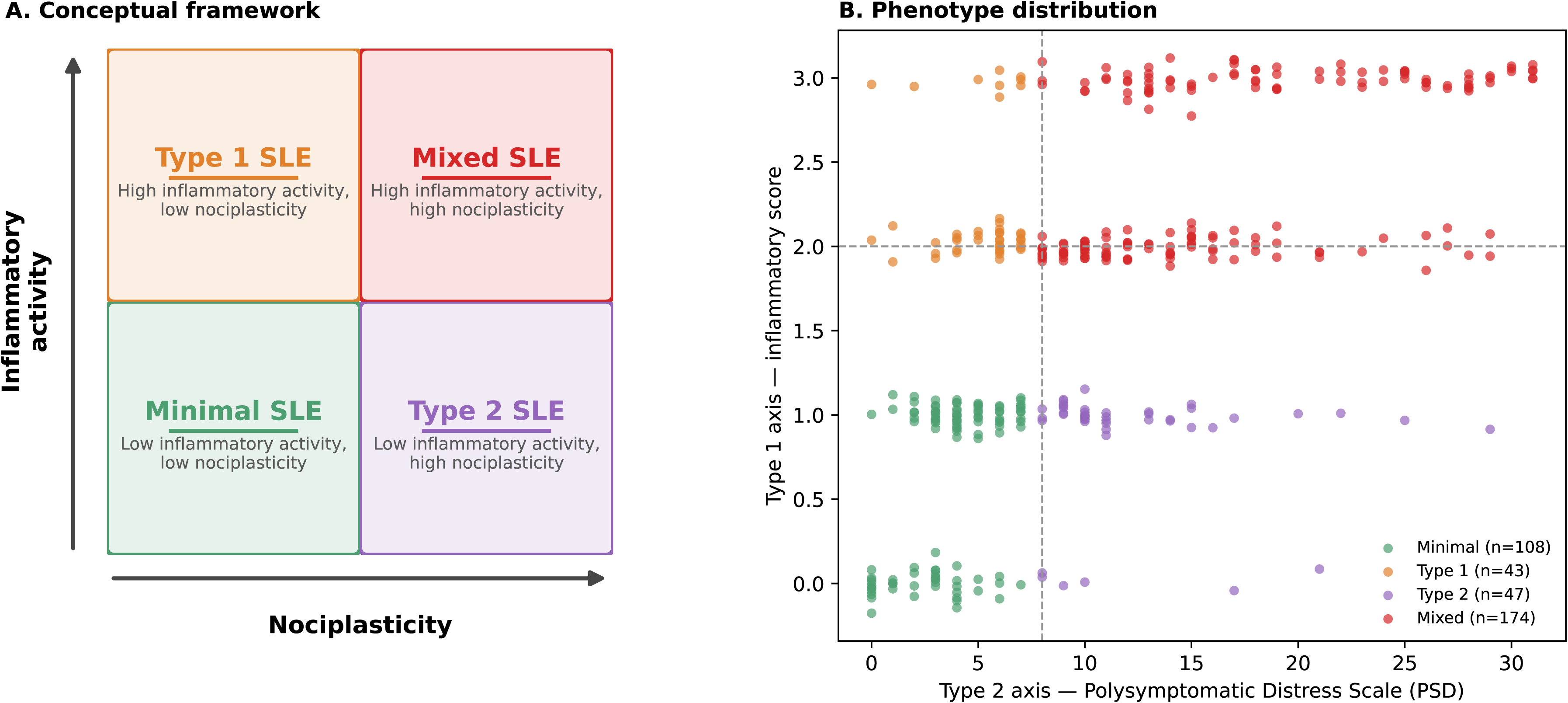
Type 1 and Type 2 SLE conceptual framework and phenotype distribution. (A) Conceptual 2×2 classification based on inflammatory activity and nociplasticity. High inflammatory activity (Type 1 axis) was defined as an inflammatory score ≥2, and high nociplasticity (Type 2 axis) was defined as a PSD score ≥8. Accordingly, patients were classified as Minimal, Type 1, Type 2, or Mixed. (B) Scatter plot of individual patients (N=372) by inflammatory score (Type 1) and PSD (Type 2); points were slightly jittered to reduce overlap; phenotype classification was based on the original values. Dashed lines indicate classification thresholds (PSD=8, inflammatory score=2).

### Outcome Measures

Pain was measured by Visual Analog Scale (VAS). Functional outcomes included Patient-Reported Outcomes Measurement Information System (PROMIS)-29 v2.0 T-scores, 36-Item Short Form Survey (SF-36) Physical and Mental Component Summaries (PCS, MCS) and subscales, the Health Assessment Questionnaire Disability Index (HAQ-DI). For PROMIS-29, higher scores indicate more of the measured construct (worse for Pain Interference, Fatigue, Sleep Disturbance, Anxiety, Depression; better for Physical Function and Satisfaction with Social Roles). For SF-36 and its subscales, higher scores indicate better health. For HAQ-DI, higher scores indicate greater disability and worse outcome. Cumulative organ damage was assessed using the Brief Index of Lupus Damage (BILD), higher scores indicate greater cumulative organ damage.

### Depression and Medication Counts

Depression was assessed using the Patient Health Questionnaire-8 (PHQ-8; score ≥10 indicating probable major depression). Medication classifications are listed in Supplementary Table 1 (rheumatology medications) and Supplementary Table 2 (non-rheumatology medications). Non-rheumatology medications were further categorized into psychiatric, pain management, gastrointestinal, cardiovascular, osteoporosis, and general categories. Logistic regression modeled opioid use as a function of inflammatory score, PSD, age and sex, to assess whether opioid prescribing tracked one or both axes; odds ratios were standardized to per standard deviation (SD) increments to permit direct comparison across the differently scaled predictors. Longitudinal opioid use was plotted as group-stratified proportions from the index visit (n=301).

### Statistical Analysis

Cross-sectional comparisons used one-way ANOVA for continuous variables and chi-squared tests for categorical variables. Functional outcomes and measurements compared across phenotype groups used ANOVA with standard error of the mean (SEM).

Longitudinal functional outcomes were assessed by plotting group-mean trajectories (binned to 1-year intervals from the index visit) for Pain VAS, SF-36 PCS, PROMIS Pain Interference, HAQ-DI, and BILD, with ≥10 observations per bin required. To test whether trajectories diverged over time, linear mixed-effects models with random intercepts and slopes were fit as outcome ∼ years × group (reference: Minimal), and a joint Wald test of the three interaction terms assessed whether slopes differed across phenotype groups.

#### Transition dynamics

Observed visit-to-visit transitions were summarized by row-normalizing the 4×4 matrix of consecutive phenotype transitions. Exit proportions (excluding retention) were compared using binomial tests.

To account for unequal intervals between visits, phenotype transitions were modeled using a continuous-time Markov model (CTMC) with transition-intensity matrix *Q*:

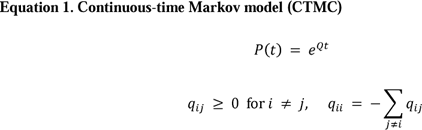

Where *P*(*t*) is the transition-probability matrix at time *t*, and *q_ij_* is the instantaneous transition rate from phenotype *i* to phenotype *j*. The model was fitted by maximum likelihood using all consecutive visit pairs and their observed time intervals. Mean sojourn time was calculated as -1/*q_ii_*, with 95% CIs from 120 patient-level bootstrap replicates. The stationary distribution was derived from *Q* as the model-implied phenotype distribution.

#### Within-patient trajectories

Population-average PSD and inflammatory score trajectories were estimated with linear mixed-effects models with random intercepts and slopes. Slopes were standardized to SD units to facilitate comparison between the two axes.

Latent trajectory subgroups were identified using a joint group-based trajectory model fitted simultaneously to PSD and inflammatory score:

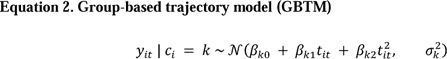

Where *y_it_* denotes either outcome for patient *i* at time *t*, and *c_i_* denotes latent class membership. Models with 2–8 classes were compared, with the final model selected using the BIC elbow criterion and a minimum class size of 5%. Within each class, patient-specific PSD and inflammatory-score slopes were estimated and tested against zero using one-sample t tests.

All analyses used complete-case analysis for each outcome. All analyses used Python with pandas, NumPy, SciPy, statsmodels libraries. Significance was set at p<0.05.

## RESULTS

### Study Cohort and Phenotype Distribution

After applying the inclusion criteria described above, 372 patients were included in the cross-sectional cohort. The cohort was 93% female, mean age 56.0 years (SD 12.8), 81% White, with mean SLE duration 21.2 years (SD 12.3). Among the 295 patients (79%) with available geographic data, patients resided in 47 US states across all four census regions (Midwest 35%, South 31%, West 21%, Northeast 13%).

Using the dual-axis classification: Type 1 (inflammatory) defined by inflammatory score ≥2, Type 2 (nociplastic) defined by PSD ≥8,^7^ 108 patients (29%) were classified as Minimal, 43 (12%) as Type 1, 47 (13%) as Type 2, and 174 (47%) as Mixed (Figure 1, Table 1). Mean PSD and inflammatory scores confirmed clear separation across groups (Supplementary Figure 2).

**Table 1.** Baseline demographic and clinical characteristics by phenotype group.

| **Characteristic** | **Overall**<br>**(N=372)** | **Minimal**<br>**(n=108)** | **Type 1**<br>**(n=43)** | **Type 2**<br>**(n=47)** | **Mixed**<br>**(n=174)** |
| --- | --- | --- | --- | --- | --- |
| Age, years | 56.0 (12.8) | 56.8 (13.8) | 55.2 (12.8) | 56.1 (13.9) | 55.6 (11.9) |
| Female | 347 (93%) | 95 (88%) | 40 (93%) | 45 (96%) | 167 (96%) |
| *Race/ethnicity* |  |  |  |  |  |
| White | 301 (81%) | 89 (82%) | 36 (84%) | 36 (77%) | 140 (80%) |
| Black | 41 (11%) | 9 (8%) | 4 (9%) | 5 (11%) | 23 (13%) |
| Other | 30 (8%) | 10 (9%) | 3 (7%) | 6 (13%) | 11 (6%) |
| *Education* |  |  |  |  |  |
| ≤High school | 10 (3%) | 0 (0%) | 1 (2%) | 1 (2%) | 8 (5%) |
| Some college | 51 (14%) | 12 (11%) | 3 (7%) | 4 (9%) | 32 (18%) |
| College or higher | 311 (84%) | 96 (89%) | 39 (91%) | 42 (89%) | 134 (77%) |
| SLE duration, years | 21.2 (12.3) | 23.8 (12.7) | 19.2 (14.5) | 20.6 (11.6) | 20.3 (11.5) |
| Inflammatory score, 0–3 | 1.7 (1.0) | 0.6 (0.5) | 2.2 (0.4) | 0.9 (0.3) | 2.5 (0.5) |
| PSD, 0–31 | 11.1 (7.9) | 3.7 (2.1) | 5.1 (2.0) | 12.3 (4.7) | 16.8 (6.8) |

*Note.* Values are mean (SD) for continuous variables and n (%) for categorical variables. SLE = systemic lupus erythematosus;
PSD = polysymptomatic distress scale, range 0–31.

### Functional Outcomes by Phenotype

Functional impairment increased monotonically from Minimal to Mixed (Figure 2): pain (VAS 1.1 to 5.8; PROMIS Pain Interference 46.2 to 63.6), physical function (SF-36 PCS 49.7 to 30.0; PROMIS Physical Function 51.5 to 37.1; HAQ-DI 0.7 to 1.1), fatigue, social participation, and mental health all worsened progressively from Minimal to Mixed (all p<0.003). BILD scores were highest in Mixed (3.9 vs 2.6–2.9).

**Figure 2.**
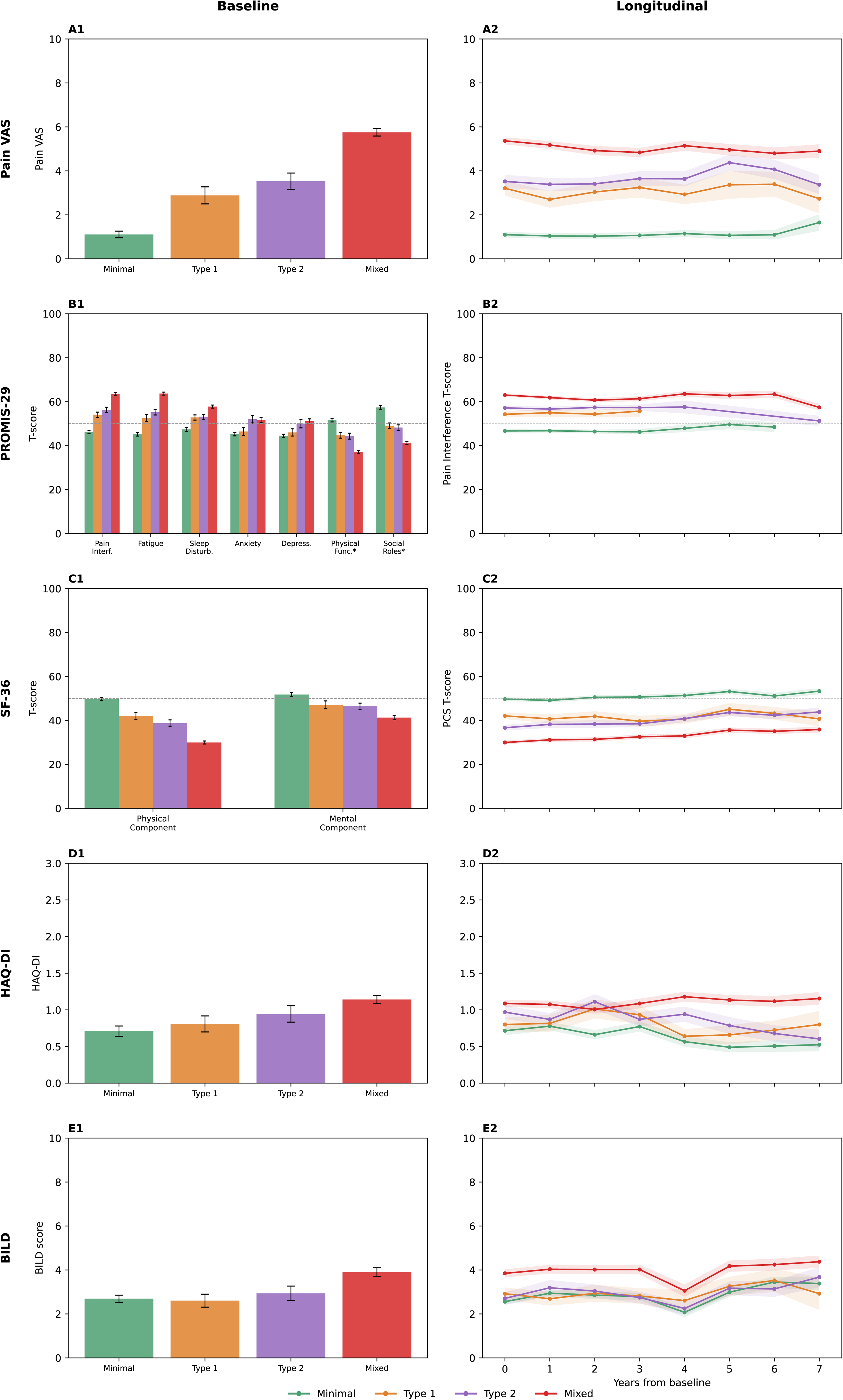
Functional outcomes by phenotype: cross-sectional (left) and longitudinal trajectories (right). (A1) Pain visual analog scale (VAS; 0–10). (A2) Pain VAS longitudinal trajectory. (B1) PROMIS-29 T-scores; asterisk (*) indicates domains in which higher scores represent better outcomes; dashed line indicates the population mean (T=50). (B2) PROMIS Pain Interference longitudinal trajectory. (C1) SF-36 Physical and Mental Component Summary scores; dashed line indicates the population mean (T=50). (C2) SF-36 Physical Component Summary (PCS) longitudinal trajectory. (D1) Health Assessment Questionnaire Disability Index (HAQ-DI; 0–3). (D2) HAQ-DI longitudinal trajectory. (E1) Brief Index of Lupus Damage (BILD; 0–10 displayed; full scale 0–47). (E2) BILD longitudinal trajectory. Shaded bands represent SEM in all longitudinal panels (A2-E2). Abbreviations: PROMIS, Patient-Reported Outcomes Measurement Information System; SF-36, 36-Item Short Form Health Survey; PCS, Physical Component Summary; HAQ-DI, Health Assessment Questionnaire Disability Index; BILD, Brief Index of Lupus Damage; SEM, standard error of the mean.

Longitudinal trajectories confirmed persistent functional separation over 7 years (Figure 2 A2– E2; n=301, median 7 visits). For Pain VAS, SF-36 PCS, and BILD, group-stratified trajectories remained parallel without significant group × time interaction. For HAQ-DI, the Mixed group worsened over time (+0.028/year, p=0.015) while the Minimal group improved (−0.038/year, p=0.006), producing progressive divergence (interaction p=0.002; Figure 2 D2). Conversely, PROMIS Pain Interference showed convergence: the Mixed group’s scores declined (−0.56 T-score/year, p<0.001) while Minimal remained flat, narrowing the between-group gap (interaction p=0.045; Figure 2 B2). BILD scores increased in all groups, consistent with progressive organ damage accumulation, but remained highest in Mixed throughout. Taken together, these findings indicate that the functional penalty of nociplastic phenotype membership persists over longitudinal follow-up; and for disability, widens.

### Depression and Medication Burden

Depression prevalence increased from 3% in Minimal to 43% in Mixed (p<0.001; Figure 3A), broadly tracking the Type 2 axis. Total medication counts likewise increased from 6.6 to 10.5 (p<0.001; Figure 3B), driven mainly by non-rheumatology medications (4.9 to 8.2; p<0.001). Psychiatric medication use rose from 24% to 64%, including SNRI use from 7% to 29% (p=0.001; Figure 3D). However, non-psychiatric medication burden also increased across groups (4.33 to 6.78; p<0.001), and substantial pain-medication use persisted among non-depressed Mixed patients (opioid 35.4% vs. 8.3% in Minimal), indicating that the excess was not attributable to psychiatric comorbidity alone.

**Figure 3.**
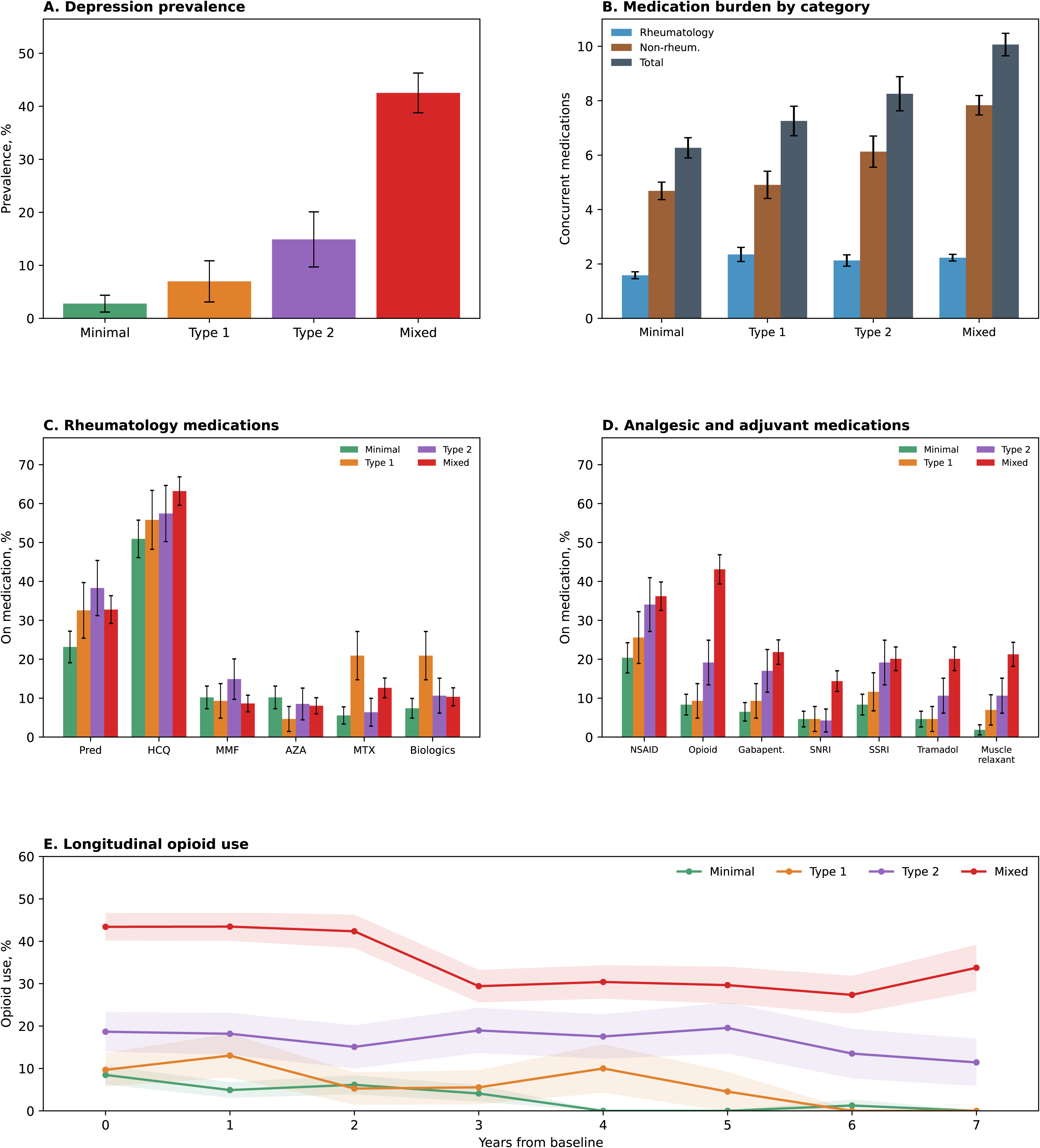
Depression and medication patterns by phenotype. (A) Depression prevalence. (B) Total medication count stratified by rheumatology vs. non-rheumatology medications. (C) Individual rheumatology medication classes (Pred, prednisone; HCQ, hydroxychloroquine; MMF, mycophenolate mofetil; AZA, azathioprine; MTX, methotrexate). (D) Non-rheumatology medication use by class, including analgesics (opioids, tramadol, muscle relaxants), neuromodulators (Gabapent., gabapentinoids), and antidepressants (SNRI, serotonin– norepinephrine reuptake inhibitors; SSRI, selective serotonin reuptake inhibitors). (E) Longitudinal opioid use (%) over up to 7 years.

Gabapentinoid use was highest in Type 2 and Mixed groups (17% and 22%), and opioid use increased from 8% in Minimal to 44% in Mixed (p<0.001; Figure 3D). Both PSD and inflammatory score were independently associated with opioid use (OR per SD 1.62, 95% CI 1.20–2.18; and 1.79, 1.28–2.51, respectively). Although opioid use declined over follow-up, it approached zero only in Minimal and Type 1, remaining 14% in Type 2 and 27–34% in Mixed by year 6-7 (Figure 3E).

### Phenotype Transitions

Among 301 patients in the longitudinal cohort, 60% experienced at least one phenotype transition (mean 1.9 per patient). From baseline to last visit, Mixed and Minimal were the most stable groups (75% and 72% retention), whereas Type 1 (15%) and Type 2 (50%) were not (Figure 4A). Visit-to-visit transition probability confirmed this pattern: Minimal and Mixed had the highest retention probabilities (0.79 and 0.78), while Type 1 and Type 2 were transient (0.33 and 0.44; Figure 4B). Critically, the two transient states had divergent exit trajectories: when Type 1 patients left their state, 49% moved to Minimal (transition probability 0.33); whereas when Type 2 patients left, 65% moved to Mixed (transition probability 0.36), with only 28% reaching Minimal (transition probability 0.16; binomial test for equal exit probability p<0.001).

**Figure 4.**
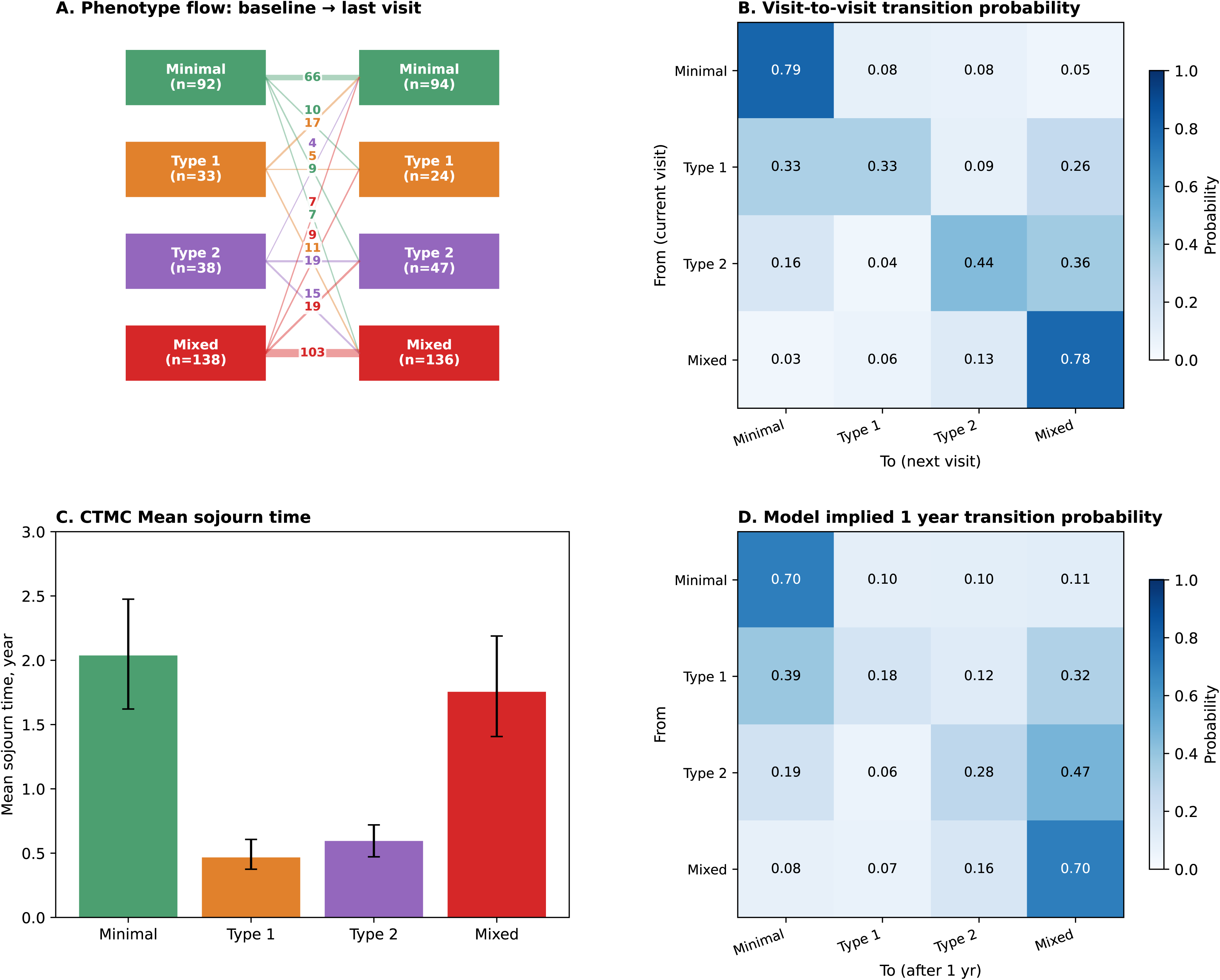
Phenotype transitions. (A) Alluvial plot showing patient flow from baseline to last visit. (B) Visit-to-visit transition probability matrix; darker shading indicates higher transition probability. Diagonal cells indicate retention within the same phenotype. (C) Mean sojourn times estimated from a continuous-time Markov chain (CTMC) model with 95% bootstrap confidence intervals. (D) Implied one-year transition probability matrix derived from the CTMC rate matrix Q via P(1) = exp(Q).

The continuous-time Markov model confirmed these dynamics (Figure 4C, 4D). Mean sojourn times (i.e., the average duration a patient remained in one phenotype before transitioning) were: Minimal 2.03 years (95% CI 1.60–2.75), Type 1 0.46 years (0.35–0.61), Type 2 0.59 years (0.47–0.77), Mixed 1.75 years (1.32–2.29; Figure 4C). The model-implied one-year transition matrix (Figure 4D) confirmed the same pattern as the observational visit-to-visit probabilities (Figure 4B): Minimal and Mixed had the highest one-year retention probability (both 0.70), while Type 1 and Type 2 were transient (0.18 and 0.28). The similarity between the model-implied stationary distribution (Minimal 32%, Type 1 9%, Type 2 15%, Mixed 44%) and observed phenotype prevalence (29%, 12%, 13%, 47%) supports the internal consistency of the transition model and suggests that the observed distribution is not driven solely by a single baseline snapshot.

### Within-Patient Trajectories

The population-average PSD trajectory was nearly flat (+0.014 SD/year, p=0.07; Figure 5A), indicating stable nociplastic burden at the population level. The inflammatory axis declined modestly (−0.034 SD/year, p=0.002; Figure 5B).

**Figure 5.**
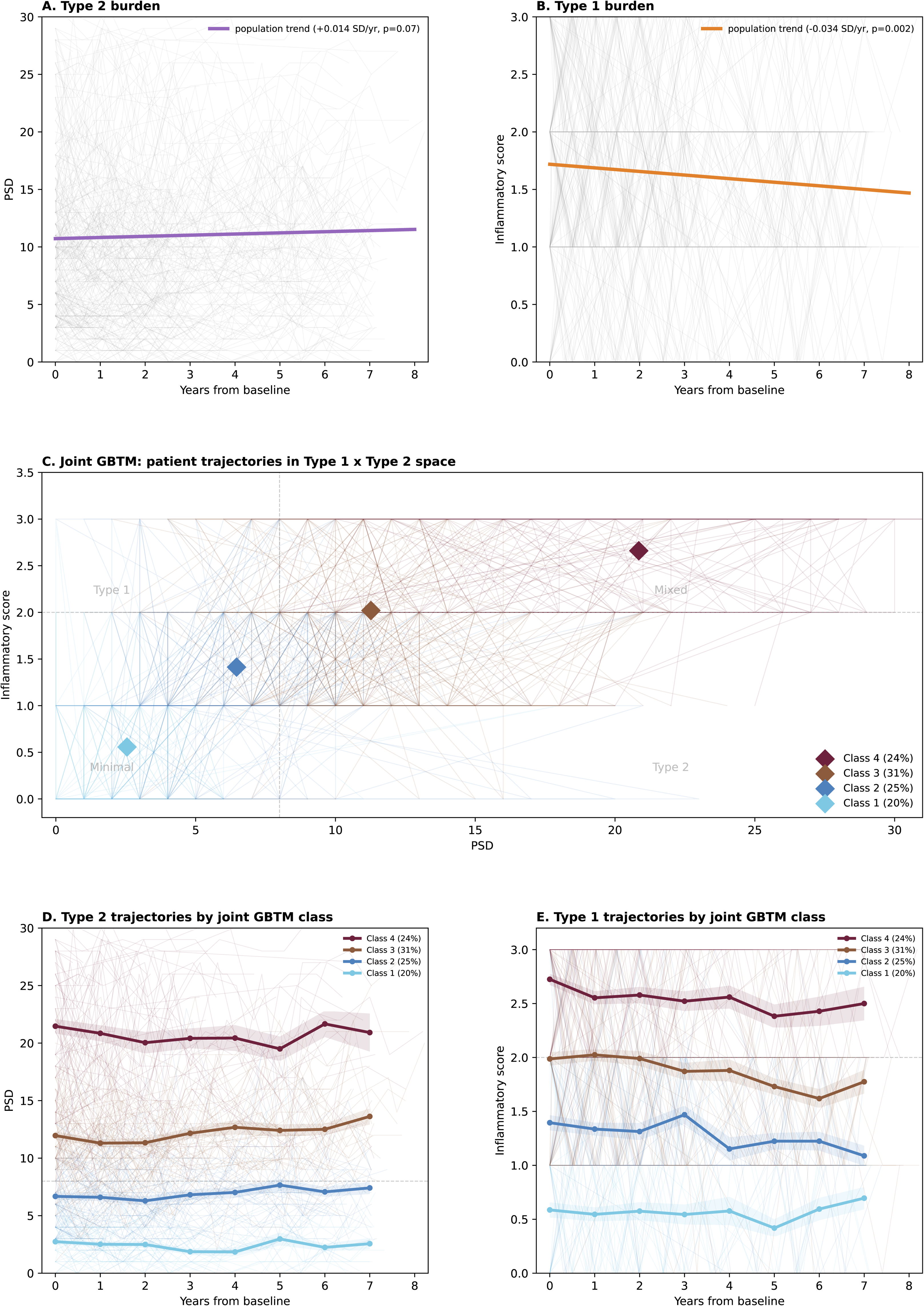
Within-patient trajectories and data-driven trajectory classes. (A) Individual PSD trajectories with population-level linear trend (+0.014 SD/year, p=0.07). Thin gray lines represent individual patient trajectories. (B) Individual inflammatory score trajectories with population-level trend (−0.034 SD/year, p=0.002). (C) Joint group-based trajectory model (GBTM) of PSD and inflammatory score (K=4 classes selected by BIC; Supplementary Figure 3). Each line represents one patient’s trajectory through PSD × inflammatory score space; diamond markers indicate class centroids. Dashed lines denote a priori classification thresholds (PSD=8, inflammatory score=2). All four data-driven classes fall along a low-to-high severity diagonal. (D) Mean PSD trajectories by GBTM class overlaid on individual trajectories. (E) Mean inflammatory score trajectories by GBTM class overlaid on individual trajectories.

To characterize the longitudinal structure of symptom burden without imposing the a priori classification, we fit a joint group-based trajectory model (GBTM) on PSD and inflammatory score simultaneously. A four-class solution was selected by BIC (Supplementary Figure 3). Plotting each patient’s longitudinal path through the PSD × inflammatory score space (Figure 5C) revealed that the four data-driven classes align along a Minimal-to-Mixed severity diagonal rather than distributing across the four quadrants of the a priori taxonomy. Class 1 (20%) remained low on both axes (mean PSD 2.5, inflammatory score 0.6). Class 2 (25%) sat below both classification thresholds (PSD 6.5, inflammatory score 1.4) with slowly declining inflammatory activity. Class 3 (31%) exceeded both thresholds (PSD 11.3, inflammatory score 2.0) with gradually rising nociplastic burden. Class 4 (24%) was persistently high on both axes (PSD 20.8, inflammatory score 2.7). All class-specific slopes were statistically indistinguishable from zero on both dimensions (Figures 5D–E; all p>0.05), confirming the longitudinal stability of symptom severity in established SLE. Notably, no stable trajectory class occupied either off-diagonal quadrant: neither Type 1 nor Type 2 emerged as durable trajectory classes. This is consistent with the transition analyses, in which Type 1 patients predominantly moved to Minimal and Type 2 patients predominantly moved to Mixed, suggesting that both off-diagonal phenotypes represent transitional states that converge toward the Minimal–Mixed severity diagonal over time.

### Sensitivity Analyses

Alternative Type 2 definitions (PSD≥11,^12^ 2011 fibromyalgia criteria,^18^ and 2016 fibromyalgia criteria)^19^ were compared with the primary PSD≥8 rule (Supplementary Figure 4A). Concordance between PSD≥8 and FM-2016 classification was 68% (Supplementary Figure 4B); stricter thresholds progressively reduced nociplastic group sizes (PSD≥8: 59%, PSD≥11: 43%, FM-2016: 27%), with discordance concentrated in the Type 2 and Mixed groups. Visit-window restriction (6±1 months) retained 83% of patients with sojourn times and trajectory slopes unchanged (Supplementary Figure 4C, 4D).

## DISCUSSION

This study provides the first longitudinal evaluation of the Type 1 and Type 2 SLE framework^7^ using patient-reported outcome (PRO)-defined phenotypes in a nationwide US registry. Three principal findings emerge: PRO-defined nociplastic phenotypes produced clinically coherent symptom gradients confirmed by stepwise functional impairment across every measured domain; the associated burden is profound and statistically stable over up to 7 years; and joint GBTM modeling on both axes reveals a single severity diagonal from Minimal to Mixed, with neither Type 1 nor Type 2 sustained as a durable latent class.

The chronicity of nociplastic symptoms is a central finding. Population-average PSD remained nearly flat over time (Figure 5A), whereas the inflammatory score declined modestly (Figure 5B). Joint GBTM of both dimensions identified four latent classes arrayed along a Minimal-to-Mixed severity diagonal (Figure 5C), rather than recovering the four a priori phenotypic quadrants. All class-specific trajectories remained essentially flat (Figure 5D–E), indicating that severity tiers remain stable once established. Likewise, no latent class centroid occupied the off-diagonal Type 1 or Type 2 quadrants, consistent with the transition analyses showing these phenotypes to be transient (Figure 4). The observed Type 1 to Minimal transitions likely reflect declining inflammatory activity crossing the predefined classification threshold rather than a fundamental change in longitudinal disease trajectory, as the underlying GBTM-derived severity trajectories remained stable across all classes. Together, these findings extend the qualitative “persistent Type 2” pattern described by Eudy et al.^10,11^ to a large, geographically diverse cohort and support the emerging concept of lupus-associated nociplasticity (LAN).^16^ Longitudinal trajectory studies in chronic widespread pain and knee osteoarthritis have similarly identified stable severity subgroups using latent class methods,^20,21^ suggesting that nociplastic stability may be a shared feature across central sensitization conditions.

The asymmetric transition dynamics carry direct clinical implications. Type 1 was the most transient phenotype (sojourn 0.46 years, Figure 4), with 49% of exits moving to Minimal, whereas Type 2 (sojourn 0.59 years) preferentially transitioned to Mixed (65% of exits). The transience of Type 1 likely reflects both regression to the mean near the inflammatory threshold and the modest population-level decline in inflammatory symptoms, whereas the stability of nociplastic burden may reflect the self-sustaining nature of central sensitization, which is less responsive to standard immunomodulatory therapy. These patterns identify a subgroup with emerging nociplastic features who warrant centrally targeted symptom-management strategies alongside inflammatory disease evaluation.

Functional outcomes revealed a stepwise increase in impairment across the nociplastic gradient on every domain. The magnitude of impairment in Mixed patients: PROMIS Pain Interference exceeding the moderate-pain threshold,^22^ SF-36 PCS comparable to advanced heart failure and end stage renal disease (Figure 2 B1-C1),^23,24^ was striking given that these patients are not typically considered candidates for pain-specialty referral. Three independent physical function measures converged on the same gradient, and the 14-fold difference in depression (3% Minimal vs 43% Mixed, Figure 3A) suggests that nociplastic burden carries psychological consequences beyond pain itself. Longitudinally, the functional penalty persisted over 7 years, and widened for disability specifically, with Mixed worsening and Minimal improving (Figure 2 A2-E2). These findings support the clinical relevance of nociplastic phenotyping beyond pain-specific instruments and underscore the call for multimodal management targeting both inflammatory and nociplastic mechanisms.^4,7,25^

The medication patterns separate into confirmatory and novel findings. As expected, depression and opioid use rose in parallel with PSD. The novel finding is that nearly half of Mixed patients were on opioids despite limited evidence for efficacy in nociplastic conditions,^26,27^ and that opioid use was independently associated with both PSD and inflammatory score, suggesting analgesic prescribing tracks both dimensions without targeted differentiation. Although claims-based studies report declining opioid use in autoimmune rheumatic diseases,^28^ our longitudinal data show that opioid prevalence in Type 2 and Mixed patients remained elevated over 7 years (Figure 3E). The 14-fold depression gradient likely drives much of the non-rheumatology medication excess through antidepressant and anxiolytic prescribing. These patterns suggest an opportunity for phenotype-guided analgesic selection to reduce opioid reliance and increase use of centrally targeted pharmacologic and non-pharmacologic strategies.^29^

We adopted the PSD≥8 threshold following recent review.^7^ Sensitivity analyses confirmed that alternative definitions (PSD≥11, FM-2016, FM-2011) yielded concordant patterns with progressively smaller nociplastic groups (Supplementary Figure 4A), consistent with nociplastic pain as a continuum^5^ in which fibromyalgia criteria capture only the severe end.^27^ The identification of patients with prominent nociplastic features may also inform evaluation of emerging biologic therapies.^30^ Future studies could use the Type 1/Type 2 framework to test whether patients with predominantly inflammatory, nociplastic, or Mixed profiles respond differently to immunomodulatory versus centrally acting interventions.

Several limitations warrant acknowledgment. The PRO-only approach cannot detect clinically silent inflammatory activity, particularly serologically active clinically quiescent (SACQ) disease or lupus nephritis, potentially misclassifying some patients. Second, because PSD and SLAQ were not available concurrently for all post-2016 patients, the analytic cohort represented a subset of eligible FORWARD patients; selection into complete PRO measurement may have enriched for patients with greater symptom burden or more consistent registry participation. Third, some associations with pain-specific outcomes are partly inherent to the PSD-based definition of Type 2 activity; therefore, the observed differences in non-pain outcomes such as physical function, social roles, depression, and medication burden are particularly important for establishing broader clinical relevance. Fourth, the FORWARD cohort is predominantly White (81%) with long-standing disease (mean 21.2 years), limiting generalizability. Although convergence with the original Duke cohort (59% Black)^8^ suggests framework transportability and highlights the need for validation in diverse and early-disease cohorts. Finally, we cannot infer clinician intent from medication data, and the observational design precludes causal inference.

In conclusion, nociplastic phenotypes are chronic, severity-stratified, and associated with profound functional impairment and high analgesic burden in a nationwide SLE cohort. Joint trajectory modeling confirms a stable severity gradient from Minimal to Mixed, with the off-diagonal phenotypes functioning as transient states: Type 1 resolving and Type 2 progressing. This evaluation extends the Type 1/Type 2 framework to community-based practice and supports integration of standardized nociplastic pain assessment into routine SLE care.

## Data Availability

Deidentified participant data are available from FORWARD, The National Databank for Rheumatic Diseases, upon reasonable request, subject to FORWARD approval and applicable data-use agreements. Analytic code is available from the corresponding author upon reasonable request.

## Contributors

CYH and TF conceived and designed the study. SP and KM provided and curated the FORWARD data. CYH and YL conducted the statistical analyses. CYH, CTS, TCD, SB, SP, KM, and TF contributed to the development and interpretation of the analytical methods. CYH created the figures and wrote the first draft of the manuscript. CTS, YL, SP, TCD, SB, PK, KM, and TF interpreted the findings and critically revised the manuscript. All authors approved the final version of the manuscript.

## Acknowledgments

This study was supported by the Lupus Research Alliance. We thank the participants of FORWARD, The National Databank for Rheumatic Diseases, whose continued participation made this study possible, and the FORWARD staff for maintaining the registry and supporting data collection.

## AI-assisted editing disclosure

During manuscript preparation, the authors used Claude (Anthropic) and ChatGPT (OpenAI) for programming assistance, language editing and internal consistency checks. The authors reviewed and revised all AI-generated output and take full responsibility for the content of the manuscript.

