## Supplementary Figures and Tables for "Longitudinal Characterization of Nociplasticity in Systemic Lupus Erythematosus: A Nationwide Registry Study"

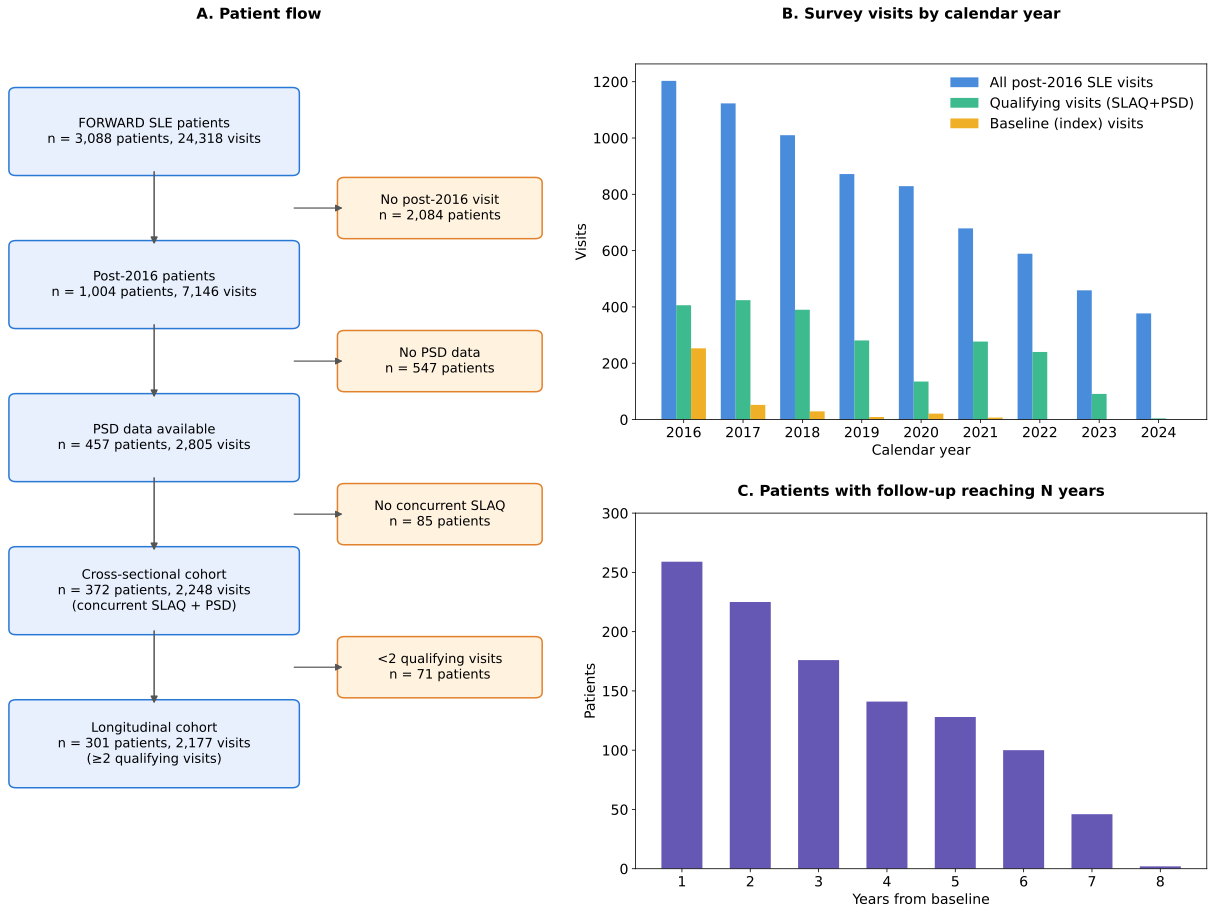

**Supplementary Figure 1.** Patient flow and data availability in the Forward SLE cohort. (A) CONSORT-style patient flow diagram showing attrition from 3,088 SLE patients to the cross-sectional (n=372) and longitudinal (n=301) analytic cohorts. (B) Number of survey visits by calendar year: all post-2016 SLE visits, qualifying visits with concurrent SLAQ and PSD data, and baseline (index) visits. (C) Number of patients with follow-up reaching each year from their baseline visit.

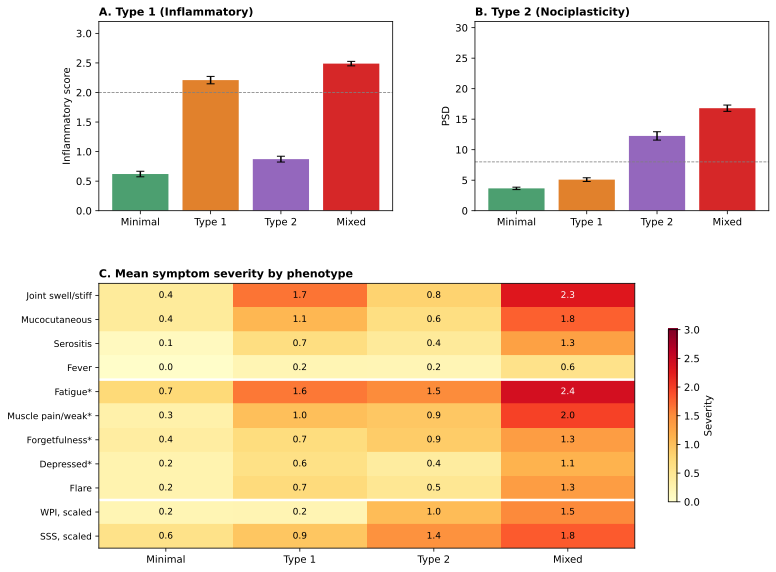

**Supplementary Figure 2.** Patient-reported outcome (PRO) severity and symptom profile by phenotype. (A) Mean inflammatory score across phenotype groups; dashed line indicates inflammatory score =2 threshold. (B) Mean PSD score; dashed line indicates PSD =8 threshold. (C) Heatmap of mean symptom severity by phenotype group; Asterisks (*) denote SLAQ symptoms excluded from the Type 1 inflammatory domains to minimize construct overlap. WPI and SSS are rescaled to 0–3 for comparability. Error bars represent SEM.

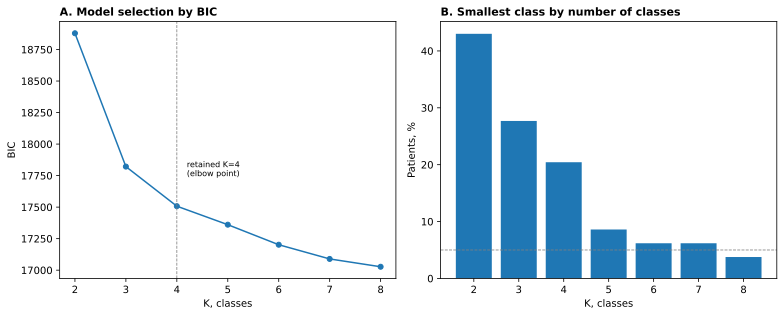

**Supplementary Figure 3.** **Joint GBTM model selection.** (A) BIC by number of classes for the joint GBTM (PSD + inflammatory score). The retained K=4 solution (dashed line) was selected at the elbow corresponding to the largest deceleration in BIC improvement. (B) Smallest class size by K; the dashed line marks the 5% minimum threshold.

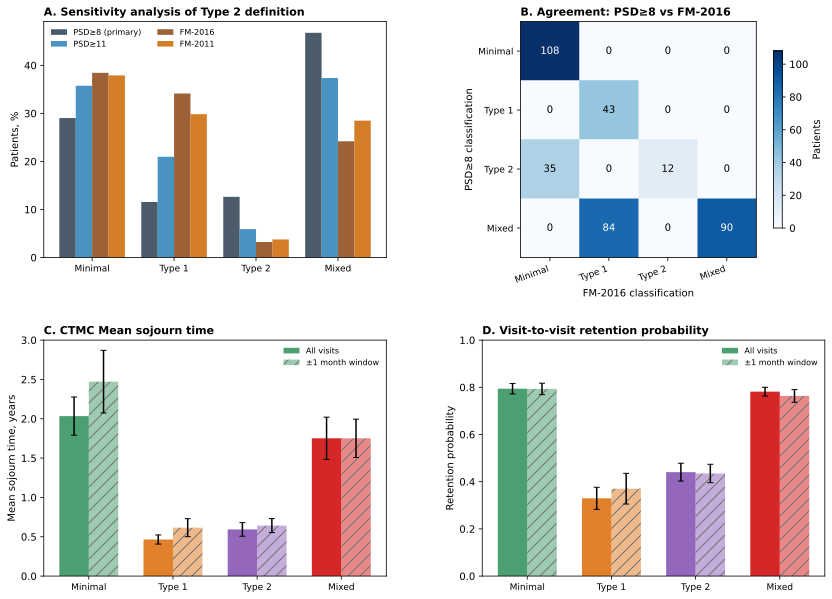

**Supplementary Figure 4. Sensitivity analyses.** (A) Phenotype distribution under four alternative Type 2 definitions. (B) Agreement matrix between PSD≥8 and FM-2016 phenotype classifications. Cell values indicate the number of patients; darker shading indicates larger patient counts. (C) Mean sojourn times estimated using all visits versus visits restricted to a 6 ± 1 month follow-up window. (D) Discrete visit-to-visit retention probabilities using all visits versus visits restricted to a 6 ± 1 month follow-up window. Error bars represent SEM.

**Supplementary Table 1.** Rheumatology medications by phenotype group. Self-reported current use of disease-modifying antirheumatic drugs (DMARDs), biologics, corticosteroids, nonsteroidal anti-inflammatory drugs (NSAIDs), and other rheumatology-related medications at the baseline qualifying visit.

| **Category** | **Generic Name** | **Minimal (n=108)** | **Type 1 (n=43)** | **Type 2 (n=47)** | **Mixed (n=174)** |
| --- | --- | --- | --- | --- | --- |
| DMARDs | Hydroxychloroquine (Plaquenil) | 70 (53%) | 47 (60%) | 12 (54%) | 87 (63%) |
| DMARDs | Methotrexate | 7 (5%) | 12 (15%) | 2 (9%) | 19 (14%) |
| DMARDs | Mycophenolate (CellCept) | 15 (11%) | 6 (8%) | 3 (14%) | 13 (9%) |
| DMARDs | Azathioprine (Imuran) | 14 (10%) | 3 (4%) | 1 (4%) | 13 (9%) |
| DMARDs | Methotrexate oral | 4 (3%) | 9 (12%) | 1 (4%) | 12 (9%) |
| DMARDs | Methotrexate injectable | 3 (2%) | 3 (4%) | 1 (4%) | 7 (5%) |
| DMARDs | Minocycline | 4 (3%) | 4 (5%) | 1 (4%) | 2 (1%) |
| DMARDs | Leflunomide (Arava) | 0 (0%) | 1 (1%) | 1 (4%) | 6 (4%) |
| DMARDs | Cyclosporine | 1 (1%) | 0 (0%) | 1 (4%) | 0 (0%) |
| DMARDs | Sulfasalazine | 0 (0%) | 1 (1%) | 1 (4%) | 0 (0%) |
| Biologics | Belimumab (Benlysta) | 4 (3%) | 9 (12%) | 0 (0%) | 8 (6%) |
| Biologics | Abatacept (Orencia) | 2 (2%) | 0 (0%) | 2 (9%) | 1 (1%) |
| Biologics | Etanercept (Enbrel) | 1 (1%) | 2 (3%) | 1 (4%) | 0 (0%) |
| Biologics | Rituximab (Rituxan) | 0 (0%) | 0 (0%) | 0 (0%) | 3 (2%) |
| Biologics | Adalimumab (Humira) | 0 (0%) | 2 (3%) | 0 (0%) | 1 (1%) |
| Biologics | Tocilizumab (Actemra) | 1 (1%) | 0 (0%) | 0 (0%) | 1 (1%) |
| Biologics | Infliximab (Remicade) | 1 (1%) | 0 (0%) | 0 (0%) | 1 (1%) |
| Biologics | Anakinra (Kineret) | 0 (0%) | 1 (1%) | 0 (0%) | 0 (0%) |
| Biologics | Certolizumab (Cimzia) | 1 (1%) | 0 (0%) | 0 (0%) | 0 (0%) |
| Corticosteroids | Prednisone | 33 (25%) | 27 (35%) | 10 (46%) | 44 (32%) |
| NSAIDs | Ibuprofen | 7 (5%) | 3 (4%) | 2 (9%) | 14 (10%) |
| NSAIDs | Meloxicam (Mobic) | 6 (4%) | 9 (12%) | 1 (4%) | 9 (6%) |
| NSAIDs | Naproxen (Naprosyn) | 6 (4%) | 5 (6%) | 3 (14%) | 9 (6%) |
| NSAIDs | Celecoxib (Celebrex) | 3 (2%) | 3 (4%) | 0 (0%) | 6 (4%) |
| NSAIDs | Diclofenac (Voltaren) | 3 (2%) | 2 (3%) | 1 (4%) | 5 (4%) |
| NSAIDs | Nabumetone (Relafen) | 1 (1%) | 3 (4%) | 1 (4%) | 5 (4%) |
| NSAIDs | Etodolac (Lodine) | 1 (1%) | 1 (1%) | 0 (0%) | 4 (3%) |
| NSAIDs | EC Aspirin (anti-inflammatory) | 4 (3%) | 0 (0%) | 0 (0%) | 1 (1%) |
| NSAIDs | Ketorolac (Toradol) | 1 (1%) | 0 (0%) | 0 (0%) | 2 (1%) |
| NSAIDs | Arthrotec | 1 (1%) | 0 (0%) | 0 (0%) | 1 (1%) |
| NSAIDs | Indomethacin (Indocin) | 0 (0%) | 0 (0%) | 0 (0%) | 1 (1%) |
| NSAIDs | Sulindac (Clinoril) | 1 (1%) | 0 (0%) | 0 (0%) | 0 (0%) |
| NSAIDs | Salsalate | 0 (0%) | 0 (0%) | 0 (0%) | 1 (1%) |
| NSAIDs | Rofecoxib (Vioxx) | 1 (1%) | 0 (0%) | 0 (0%) | 0 (0%) |
| NSAIDs | Asatyl | 0 (0%) | 1 (1%) | 0 (0%) | 0 (0%) |
| Other Rheumatology | Folic acid | 20 (15%) | 14 (18%) | 4 (18%) | 24 (17%) |
| Other Rheumatology | Glucosamine | 4 (3%) | 5 (6%) | 0 (0%) | 4 (3%) |
| Other Rheumatology | Gout medications | 1 (1%) | 5 (6%) | 0 (0%) | 4 (3%) |
| Other Rheumatology | Herbal arthritis medications | 0 (0%) | 1 (1%) | 0 (0%) | 1 (1%) |
| Other Rheumatology | Camoquin (Chloroquine) | 1 (1%) | 0 (0%) | 0 (0%) | 0 (0%) |
| Other Rheumatology | Psoriasis medications | 0 (0%) | 0 (0%) | 0 (0%) | 1 (1%) |

**Supplementary Table 2.** Non-rheumatology medications by phenotype group. Self-reported current use of analgesics/opioids, antidepressants, anxiolytics/sedatives, anticonvulsants, muscle relaxants, and other non-rheumatology medications at the baseline qualifying visit.

| **Category** | **Generic Name** | **Minimal (n=108)** | **Type 1 (n=43)** | **Type 2 (n=47)** | **Mixed (n=174)** |
| --- | --- | --- | --- | --- | --- |
| Analgesics / Opioids | Tramadol | 5 (5%) | 2 (5%) | 5 (11%) | 35 (20%) |
| Analgesics / Opioids | Tylenol+Hydrocodone | 4 (4%) | 2 (5%) | 2 (4%) | 29 (17%) |
| Analgesics / Opioids | Other pain medications | 1 (1%) | 0 (0%) | 0 (0%) | 8 (5%) |
| Analgesics / Opioids | Morphine | 0 (0%) | 0 (0%) | 1 (2%) | 7 (4%) |
| Analgesics / Opioids | Tylenol+Oxycodone | 0 (0%) | 0 (0%) | 0 (0%) | 7 (4%) |
| Analgesics / Opioids | Oxycodone | 0 (0%) | 0 (0%) | 0 (0%) | 5 (3%) |
| Analgesics / Opioids | Tylenol+Butalbital | 1 (1%) | 0 (0%) | 0 (0%) | 3 (2%) |
| Analgesics / Opioids | Tylenol+Codeine | 0 (0%) | 0 (0%) | 1 (2%) | 3 (2%) |
| Analgesics / Opioids | Fentanyl | 0 (0%) | 0 (0%) | 0 (0%) | 3 (2%) |
| Analgesics / Opioids | Peripheral nerve block | 0 (0%) | 0 (0%) | 0 (0%) | 3 (2%) |
| Analgesics / Opioids | Codeine | 0 (0%) | 0 (0%) | 0 (0%) | 1 (1%) |
| Analgesics / Opioids | Oxymorphone | 0 (0%) | 0 (0%) | 0 (0%) | 1 (1%) |
| Analgesics / Opioids | Other strong narcotics | 0 (0%) | 0 (0%) | 0 (0%) | 1 (1%) |
| Analgesics / Opioids | Tylenol+Tramadol | 0 (0%) | 1 (2%) | 0 (0%) | 0 (0%) |
| Antidepressants | SSRI | 9 (8%) | 5 (12%) | 9 (19%) | 35 (20%) |
| Antidepressants | SNRI (other) | 4 (4%) | 3 (7%) | 6 (13%) | 34 (20%) |
| Antidepressants | Duloxetine (Cymbalta) | 5 (5%) | 2 (5%) | 2 (4%) | 23 (13%) |
| Antidepressants | Amitriptyline | 4 (4%) | 1 (2%) | 1 (2%) | 14 (8%) |
| Antidepressants | Venlafaxine (Effexor) | 4 (4%) | 3 (7%) | 1 (2%) | 7 (4%) |
| Antidepressants | Nortriptyline | 1 (1%) | 0 (0%) | 1 (2%) | 5 (3%) |
| Antidepressants | Doxepin | 0 (0%) | 1 (2%) | 1 (2%) | 1 (1%) |
| Antidepressants | Milnacipran (Savella) | 0 (0%) | 0 (0%) | 0 (0%) | 2 (1%) |
| Antidepressants | Desipramine | 0 (0%) | 0 (0%) | 0 (0%) | 1 (1%) |
| Antidepressants | Imipramine | 0 (0%) | 0 (0%) | 0 (0%) | 1 (1%) |
| Cardiovascular | Low-dose aspirin | 39 (36%) | 10 (23%) | 12 (26%) | 49 (28%) |
| Cardiovascular | Other BP medications | 22 (20%) | 14 (33%) | 16 (34%) | 51 (29%) |
| Cardiovascular | Other heart medications | 12 (11%) | 4 (9%) | 7 (15%) | 54 (31%) |
| Cardiovascular | Diuretics | 14 (13%) | 6 (14%) | 9 (19%) | 45 (26%) |
| Cardiovascular | ACE inhibitors | 19 (18%) | 5 (12%) | 10 (21%) | 25 (14%) |
| Cardiovascular | ARBs | 12 (11%) | 4 (9%) | 5 (11%) | 26 (15%) |
| Cardiovascular | Atorvastatin (Lipitor) | 12 (11%) | 2 (5%) | 7 (15%) | 12 (7%) |
| Cardiovascular | Warfarin (Coumadin) | 9 (8%) | 1 (2%) | 2 (4%) | 16 (9%) |
| Cardiovascular | Simvastatin (Zocor) | 6 (6%) | 3 (7%) | 4 (8%) | 11 (6%) |
| Cardiovascular | Pravastatin (Pravachol) | 1 (1%) | 2 (5%) | 2 (4%) | 6 (3%) |
| Cardiovascular | Rosuvastatin (Crestor) | 5 (5%) | 1 (2%) | 1 (2%) | 3 (2%) |
| Cardiovascular | Clopidogrel (Plavix) | 1 (1%) | 0 (0%) | 2 (4%) | 4 (2%) |
| Cardiovascular | Fenofibrate (TriCor) | 1 (1%) | 2 (5%) | 0 (0%) | 2 (1%) |
| Cardiovascular | Niacin | 0 (0%) | 0 (0%) | 0 (0%) | 3 (2%) |
| Cardiovascular | Rivaroxaban (Xarelto) | 1 (1%) | 0 (0%) | 0 (0%) | 2 (1%) |
| Cardiovascular | Colesevelam (Welchol) | 1 (1%) | 0 (0%) | 0 (0%) | 2 (1%) |
| Cardiovascular | Ezetimibe (Zetia) | 1 (1%) | 1 (2%) | 1 (2%) | 0 (0%) |
| Cardiovascular | Enoxaparin (Lovenox) | 0 (0%) | 0 (0%) | 0 (0%) | 2 (1%) |
| Cardiovascular | Lovastatin (Mevacor) | 1 (1%) | 0 (0%) | 0 (0%) | 1 (1%) |
| Cardiovascular | Other cholesterol medications | 0 (0%) | 1 (2%) | 0 (0%) | 0 (0%) |
| Cardiovascular | Colestipol | 1 (1%) | 0 (0%) | 0 (0%) | 0 (0%) |
| Cardiovascular | Cholestyramine (Questran) | 0 (0%) | 0 (0%) | 0 (0%) | 1 (1%) |
| Cardiovascular | Icosapent ethyl (Vascepa) | 0 (0%) | 0 (0%) | 0 (0%) | 1 (1%) |
| GI Medications | Other GI medications | 13 (12%) | 5 (12%) | 8 (17%) | 44 (25%) |
| GI Medications | Omeprazole (Prilosec) | 13 (12%) | 5 (12%) | 5 (11%) | 46 (26%) |
| GI Medications | Pantoprazole (Protonix) | 6 (6%) | 7 (16%) | 7 (15%) | 16 (9%) |
| GI Medications | Ranitidine (Zantac) | 4 (4%) | 0 (0%) | 1 (2%) | 13 (8%) |
| GI Medications | Antacids | 5 (5%) | 0 (0%) | 4 (8%) | 6 (3%) |
| GI Medications | Esomeprazole (Nexium) | 2 (2%) | 2 (5%) | 2 (4%) | 9 (5%) |
| GI Medications | Famotidine (Pepcid) | 1 (1%) | 2 (5%) | 0 (0%) | 6 (3%) |
| GI Medications | Sucralfate (Carafate) | 0 (0%) | 0 (0%) | 0 (0%) | 6 (3%) |
| GI Medications | Lansoprazole (Prevacid) | 0 (0%) | 1 (2%) | 0 (0%) | 5 (3%) |
| GI Medications | Rabeprazole | 1 (1%) | 0 (0%) | 1 (2%) | 1 (1%) |
| GI Medications | Cimetidine (Tagamet) | 0 (0%) | 0 (0%) | 0 (0%) | 1 (1%) |
| General / Other | Vitamins/supplements | 73 (68%) | 23 (54%) | 36 (77%) | 104 (60%) |
| General / Other | Calcium supplements | 47 (44%) | 10 (23%) | 15 (32%) | 44 (25%) |
| General / Other | Pulmonary medications | 14 (13%) | 14 (33%) | 10 (21%) | 73 (42%) |
| General / Other | Thyroid medications | 28 (26%) | 13 (30%) | 15 (32%) | 54 (31%) |
| General / Other | Miscellaneous | 12 (11%) | 5 (12%) | 7 (15%) | 51 (29%) |
| General / Other | Eye medications | 14 (13%) | 7 (16%) | 12 (26%) | 36 (21%) |
| General / Other | Topical medications | 9 (8%) | 5 (12%) | 6 (13%) | 30 (17%) |
| General / Other | Antibiotics | 10 (9%) | 4 (9%) | 6 (13%) | 21 (12%) |
| General / Other | Estrogen/HRT | 7 (6%) | 6 (14%) | 3 (6%) | 25 (14%) |
| General / Other | Diabetes medications | 7 (6%) | 4 (9%) | 2 (4%) | 15 (9%) |
| General / Other | Oral contraceptives | 2 (2%) | 2 (5%) | 4 (8%) | 8 (5%) |
| General / Other | Cancer medications | 3 (3%) | 1 (2%) | 0 (0%) | 7 (4%) |
| Neuromodulators | Gabapentin | 4 (4%) | 3 (7%) | 8 (17%) | 29 (17%) |
| Neuromodulators | Other anticonvulsants | 4 (4%) | 2 (5%) | 3 (6%) | 24 (14%) |
| Neuromodulators | Pregabalin (Lyrica) | 3 (3%) | 1 (2%) | 0 (0%) | 11 (6%) |
| Neuromodulators | Clonazepam | 3 (3%) | 2 (5%) | 1 (2%) | 5 (3%) |
| Neuromodulators | Carbamazepine | 0 (0%) | 0 (0%) | 1 (2%) | 3 (2%) |
| Neuromodulators | Valproic acid | 0 (0%) | 0 (0%) | 0 (0%) | 2 (1%) |
| Neuromodulators | Phenytoin | 0 (0%) | 0 (0%) | 1 (2%) | 0 (0%) |
| Osteoporosis | Alendronate (Fosamax) | 3 (3%) | 0 (0%) | 1 (2%) | 6 (3%) |
| Osteoporosis | Other osteoporosis | 4 (4%) | 0 (0%) | 1 (2%) | 2 (1%) |
| Osteoporosis | Teriparatide (Forteo) | 0 (0%) | 0 (0%) | 2 (4%) | 1 (1%) |
| Osteoporosis | Risedronate (Actonel) | 1 (1%) | 0 (0%) | 1 (2%) | 0 (0%) |
| Osteoporosis | Ibandronate (Boniva) | 1 (1%) | 0 (0%) | 0 (0%) | 1 (1%) |
| Osteoporosis | Raloxifene (Evista) | 2 (2%) | 0 (0%) | 0 (0%) | 0 (0%) |
| Osteoporosis | Bisphosphonates (other) | 1 (1%) | 0 (0%) | 0 (0%) | 1 (1%) |
| Other CNS (sleep/anxiety/muscle relaxant) | Muscle relaxants | 2 (2%) | 3 (7%) | 5 (11%) | 37 (21%) |
| Other CNS (sleep/anxiety/muscle relaxant) | Sleep medications | 6 (6%) | 4 (9%) | 5 (11%) | 31 (18%) |
| Other CNS (sleep/anxiety/muscle relaxant) | Anxiolytics | 5 (5%) | 3 (7%) | 7 (15%) | 30 (17%) |
| Other CNS (sleep/anxiety/muscle relaxant) | Migraine medications | 0 (0%) | 1 (2%) | 0 (0%) | 14 (8%) |
